# Insulin resistance assessed by short insulin tolerance test and its association with obesity and insulin resistance-related parameters in humans: A randomized trial

**DOI:** 10.1101/2024.01.12.24301240

**Authors:** Akiko Hayashishita, Taku Watanabe, Naoko Suzuki, Toshitaka Nakaya, Ayako Sugimoto, Isao Yokota, Hiroshi Ohira, Masaharu Nishimura, Ichizo Tsujino

## Abstract

The aim of this study was to examine the association of insulin resistance (evaluated by the short insulin tolerance test [SITT]) with parameters related to obesity and insulin resistance. We prospectively recruited controls and patients with type 2 diabetes mellitus (T2DM), subjected them to the SITT, and calculated the K indices of the intravenous insulin tolerance test (K_ITT_(iv)) and the subcutaneous insulin tolerance test (K_ITT_(sc)). We compared K_ITT_(iv) between the volunteers and patients, and examined its correlation with K_ITT_(sc). We also examined the association of K_ITT_(iv) with obesity, insulin resistance-related parameters, and the insulin dose required for glycemic control. A total of 24 participants (seven controls and 17 patients with T2DM) were studied. The mean K_ITT_(iv) was significantly lower in patients with T2DM than in the controls (2.5%±2.1% vs. 4.5%±1.8%). In all participants, K_ITT_(iv) was significantly correlated with the HOMA-IR values (r=−0.601, p<0.05) but not with K_ITT_(sc) (p=0.62). K_ITT_(iv) was correlated positively with the serum adiponectin concentration, but negatively with the visceral fat area and serum concentrations of tumor necrosis factor-α and branched-chain amino acids. In patients with T2DM, K_ITT_(iv) and HOMA-IR values were significantly correlated with the total insulin dose required for glycemic control. Insulin resistance evaluated using intravenous insulin (K_ITT_(iv)) was correlated with the HOMA-IR values, but not with the resistance evaluated using subcutaneous insulin (K_ITT_(sc)). The degree of insulin resistance was associated with biomarkers, such as adiponectin, and with the dose of insulin required for glycemic control.

## Introduction

Insulin resistance is a cardinal feature of the pathogenesis of non-insulin-dependent diabetes mellitus [1]. Precise estimation of insulin resistance is important to understand the mechanism underlying the impairment of glycemic control and accordingly optimize the treatment for patients with diabetes.

The hyperinsulinemic euglycemic glucose clamp test (HEGCT) is the gold standard for evaluating insulin resistance [2]. However, it is complicated and difficult to perform in many patients. The homeostasis model assessment for insulin resistance (HOMA-IR value) is another test that is frequently used for evaluating the insulin resistance. HOMA-IR can be performed readily and is well correlated with the HEGCT [3]. However, in patients with diabetes mellitus with high fasting plasma glucose (FPG) levels (who may have a decreased insulin secretion), it is less accurate as an indicator of insulin resistance [4]. In addition, HOMA-IR cannot estimate insulin resistance in patients with diabetes who are receiving insulin therapy [4].

The short insulin tolerance test (SITT) is a simple and convenient method for estimating insulin resistance [5, 6]. In the SITT, regular insulin is administered intravenously, and blood samples are collected sequentially for 15 min; the plasma glucose disappearance rate is then estimated [5, 6]. The K index of the insulin tolerance test (K_ITT_) is calculated from the linear slope of the plasma glucose concentration curve and correlates well with the HEGCT [3, 7]. The SITT can assess whole-body insulin sensitivity even in patients with decreased insulin secretion or on insulin therapy. However, the SITT carries a risk of hypoglycemia [5, 8], and K_ITT_ estimation requires multiple blood sampling. Theoretically, K_ITT_ can be estimated more easily by administering insulin subcutaneously without venous blood sampling, e.g., finger pricking, and using the glucose concentration in finger-prick blood. Furthermore, rapid hypoglycemia is less likely to occur with subcutaneous insulin administration than with intravenous insulin administration. However, the accuracy of K_ITT_ has not been validated.

Previous studies have provided the K_ITT_ values from intravenous SITT (K_ITT_(iv)) in humans [3, 5, 6, 7, 9]. Especially in patients with poorly controlled diabetes mellitus, K_ITT_(iv) is considered more accurate than HOMA-IR because it is not affected by a decreased insulin secretory capacity. However, few studies [10] have examined the association between K_ITT_(iv) and other parameters related to obesity and insulin resistance (such as visceral fat, adiponectin, and tumor necrosis factor [TNF]-α). Such data will improve the understanding of the pathogenesis of insulin resistance and help optimize the treatment in each patient. Furthermore, the application of K_ITT_(iv) for predicting the amount of insulin required would be useful in practice.

In the present prospective study, we calculated K_ITT_(iv) in healthy volunteers and patients with type 2 diabetes mellitus (T2DM). Using KITT(iv) as a reference, we then evaluated the accuracy of HOMA-IR and the K_ITT_ obtained using glucose concentration of fingertip blood (i.e., after subcutaneous insulin injection [KITT(sc)]). We also investigated the relationship between K_ITT_(iv) and the parameters related to obesity and insulin resistance, and examined the possible role of K_ITT_(iv) in predicting the insulin dose required for glycemic control in patients with T2DM.

## Methods

### Study participants

This was a single-center, randomized, crossover study with 17 patients with T2DM (who were admitted to the Hokkaido University Hospital for glycemic control) and seven healthy volunteers (controls). The registration period was from November 2016 to July 2019.

The inclusion criteria for the controls were as follows: age ≥30 years; body mass index, >18.5 kg/m^2^; and no previous diagnosis of diabetes or impaired glucose tolerance. The inclusion criteria for the patients were as follows: age ≥ 20 years, T2DM diagnosis based on the American Diabetes Association Criteria [11], and FPG level ≥ 140 mg/dL during hospitalization. The exclusion criteria for the controls were as follows: history of hypoglycemia (blood glucose < 60 mg/dL), untreated ischemic heart disease, epilepsy, pregnancy, possible pregnancy, lactation, and researcher-determined ineligibility. The exclusion criteria for the patients were as follows: unstable diabetic retinopathy, stage ≥4 diabetic nephropathy, untreated ischemic heart disease, history of hypersensitivity to human regular insulin, epilepsy, pregnancy, possible pregnancy, lactation, and deemed ineligible by the researcher.

### Crossover assignment to the insulin tolerance test with intravenous or subcutaneous insulin

After case enrollment using the central registration system by the research persons, the participants were assigned to undergo either of the following pathways: 1) the SITT first (with intravenous insulin injection), followed by an insulin tolerance test with subcutaneous insulin injection (ITTsc) or 2) the ITTsc first, followed by the SITT. This assigning of the patients was performed on a 1:1 basis by the research office, and the results were communicated to the co-investigators in writing. The two insulin tolerance tests (SITT and ITTsc) were completed within 3 days, irrespective of the order(Fig 1).

**Fig 1.**
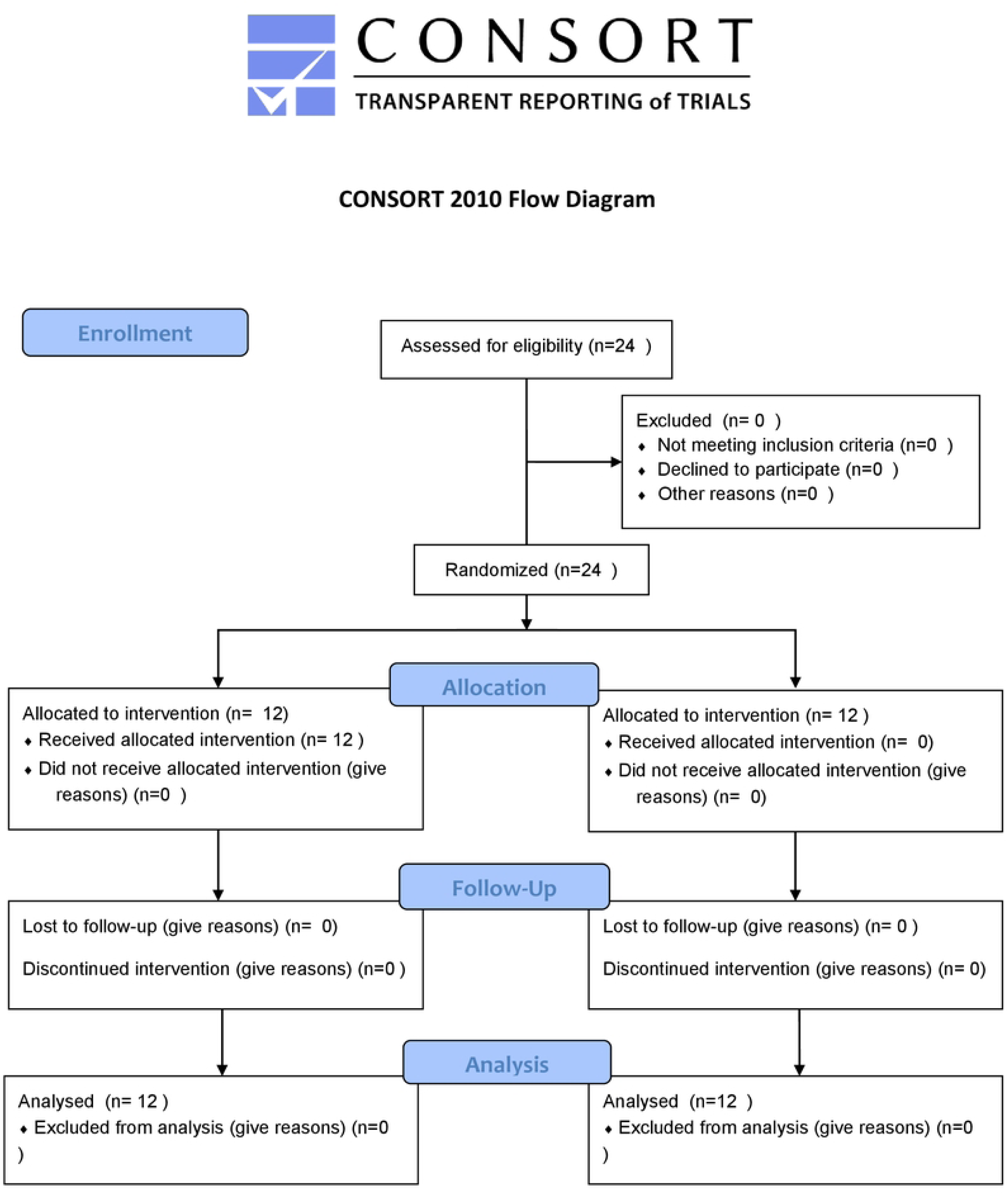

### Pre-test preparations

The controls arrived at the hospital in the morning after fasting overnight to participate in the study. Patients with T2DM, after admission to the hospital, were started on a diet (25–30 kcal/kg standard body weight/day); metformin, pioglitazone, and sodium glucose transporter 2 inhibitors were withdrawn on admission. The patients’ fasting venous blood was collected the day after admission; if the FPG level was >140 mg/dL, the patient was included in this study. The controls and patients both underwent the SITT and ITTsc after overnight fasting.

### SITT procedures

The SITT was performed before breakfast. Venous blood samples were collected for measurement of plasma glucose before and at 3, 6, 9, 12, and 15 min after an intravenous bolus injection of regular human insulin (0.1 U/kg bodyweight; Eli Lilly®, Eli Lilly and Company, IN, USA). Fifteen minutes after the insulin injection, the test was completed by injecting 20 mL of 50% glucose solution. K_ITT_ [now labelled K_ITT_(iv)] was calculated from the linear slope of the curve of plasma glucose concentrations at the 3–15-min timepoints using the Lundbaek equation [12].

### ITTsc procedures

The ITTsc was performed before breakfast. Finger-prick blood samples were collected for the measurement of blood glucose before, and at 3, 6, 9, 12, 15, 30, 45, 60, 75, 90, and 120 min after a subcutaneous injection of regular human insulin (0.1 U/kg bodyweight). The K_ITT_ (now labelled K_ITT_(sc)) was calculated from the linear slope of the curve of finger-pick blood glucose concentrations at the 30-120 min timepoints using the Lundbaek equation.

### Clinical parameters related to insulin resistance

The waist circumference and waist-to-hip ratio before breakfast were measured during the first ITT. The visceral fat area (VFA) was calculated using the bioimpedance method (BIM; InBody770^®^, InBody Japan Inc., Tokyo, Japan). We measured the fasting serum C-peptide immunoreactivity (CPR); levels of fasting immunoreactive insulin (F-IRI), adiponectin, leptin, and TNF-α; and concentrations of plasma amino acids (AA) and branched chain AAs (BCAA) using the baseline (0 min) blood sample from the SITT or ITTsc (whichever was performed first). The HOMA-IR values were calculated using the following formula: HOMA-IR = (FPG [mg/dL] × F-IRI [μU/mL])/405. F-IRI and the HOMA-IR values were not determined in insulin-treated patients.

### Insulin treatment after SITT/ITTsc

After the SITT and ITTsc, all 17 patients with T2DM were treated with multiple daily insulin injections. Basal and bolus insulin doses were adjusted until the FPG levels were ≤130 mg/dL and the postprandial plasma glucose levels were ≤180 mg/dL. The total insulin dose used for glycemic control was then calculated. Using K_ITT_(iv) and the HOMA-IR value, equations by which the total insulin dose (/body weight) could be calculated were formed.

### Statistical analyses

Data are expressed as mean ± standard deviation. The required sample size was determined to be 19 after assuming a correlation coefficient of 0.6 between K_ITT_(iv) and K_ITT_(sc), a power of 80%, and a significance level of 5%. However, the actual sample size was set at 24, assuming a 20% dropout rate.

We compared K_ITT_(iv) between the controls and patients with T2DM. We then examined the correlation of K_ITT_(iv) with the HOMA-IR value and K_ITT_(sc) using Pearson’s correlation coefficient analysis. The intraclass correlation coefficient (ICC) was also used to assess the relationship between K_ITT_(iv) and K_ITT_(sc). We further analyzed the association of K_ITT_(iv) with VFA and other parameters related to obesity and insulin resistance using a Pearson’s correlation coefficient analysis. In addition, the association of K_ITT_(iv) and the HOMA-IR value with the daily insulin dose required for glycemic control was evaluated in patients with diabetes, using Pearson’s correlation coefficient analysis. The frequency of adverse events was analyzed using the McNemar test. All analyses were performed using JMP (SAS institute, Cary, NC, USA).

Written informed consent was obtained from all individuals prior to participation in the study. This study was approved by the ethics committee of the Hokkaido University Hospital (date of approval 25 October 2016; approval no. 016-0014) and registered in the University Hospital Medical Information Network (UMIN000024453).

## Results

The clinical characteristics of the participants are summarized in Table 1. Among the 17 patients with T2DM, eight (47.1%) were treated with antihypertensive agents, eight (47.1%) with hyperlipidemic agents, two (11.8%) with sulfonylurea, five (29.4%) with insulin, six (35.3%) with metformin, five (29.4%) with dipeptidyl peptidase 4 inhibitors, and two (11.8%) with sodium glucose cotransporter 2 inhibitors.

**Table 1.** Characteristics of the study participants.

|  | Healthy controls | Patients with type 2 diabetes mellitus | p values |
| --- | --- | --- | --- |
| n (men/women) | 7 (4/3) | 17 (14/3) |  |
| Age (years) | 39.3±13.3 | 62.2±23.0 | <0.05 |
| Body weight (kg) | 62.7±18.3 | 68.5±19.1 | 0.19 |
| Body mass index (kg/m<sup>2</sup>) | 22.3±4.8 | 24.9±5.2 | <0.05 |
| Waist circumference (cm) | 78.4±12.7 | 93.6±16.4 | <0.05 |
| Waist-to-hip ratio | 0.8±0.1 | 1.0±0.1 | <0.05 |
| Visceral fat area (cm<sup>2</sup>) | 55.5±33.9 | 92.8±63.0 | <0.05 |
| Fasting plasma glucose (mg/dL) | 94.0±15.2 | 172.1±93.6 | <0.05 |
| Immunoreactive insulin (μIU/mL) | 4.7±3.7 | 4.5±4.3 | 0.67 |
| Fasting C-peptide radioactivity (ng/mL) | 1.2±0.6 | 1.7±1.0 | <0.05 |
| HOMA-IR | 1.1±0.9<sup>a</sup> | 1.9±2.3<sup>b</sup> | 0.09 |
| K<sub>ITT(iv)</sub> (%) | 4.5±1.8 | 2.5±2.1 | <0.05 |
| K<sub>ITT(sc)</sub> (%) | 0.3±0.3 | 0.5±0.5 | <0.05 |
| Adiponectin (μg/mL) | 10.9±10.1 | 5.9±4.0 | <0.05 |
| Leptin (ng/mL) | 8.1±6.7 | 10.0±8.6 | 0.31 |
| Adiponectin/Leptin | 1.5±1.2 | 0.7±1.1 | <0.05 |
| Tumor necrosis factor-α (pg/mL) | 0.6±0.9 | 1.2±0.5 | <0.05 |
| Branched chain amino acid/total amino acid | 0.15±0.04 | 0.16±0.03 | <0.05 |

<sup>a</sup> n=6
<sup>b</sup> n=11
HOMA-IR, homeostatic model assessment for insulin resistance; K<sub>ITT(iv)</sub>, K index of the intravenous insulin tolerance test; K<sub>ITT(sc)</sub> calculated from the linear slope of the curve of plasma glucose concentrations at the 30–120-min timepoints using the Lundbaek equation

The mean K_ITT_(iv) was significantly higher in the controls than in the patients (4.5%±1.8% vs. 2.%5±2.1%). Conversely, the mean HOMA-IR value was lower in the controls than in the patients (1.1±0.9 vs. 1.9±2.3). However, the mean K_ITT_(sc) did not differ significantly between the controls and the patients (0.3%±0.3% vs. 0.5%±0.5%).

In all participants, there was a significant correlation between K_ITT_(iv) and the HOMA-IR value (r=−0.601, p<0.05; Fig 2a); conversely, there was no significant correlation between K_ITT_(iv) and K_ITT_(sc) (ICC=0, p=0.62; Fig 2b). As shown in Fig 3, K_ITT_(iv) was negatively correlated with FPG (r=−0.673, p<0.05), CPR (r=−0.489, p<0.05), TNF-α (r=−0.648, p<0.05), BCAA/total AA (r=−0.64, p<0.05), and VFA (BIM; r=−0.444, p<0.05); it was positively correlated with the adiponectin concentration (r=0.487, p<0.05).

**Fig 2.**
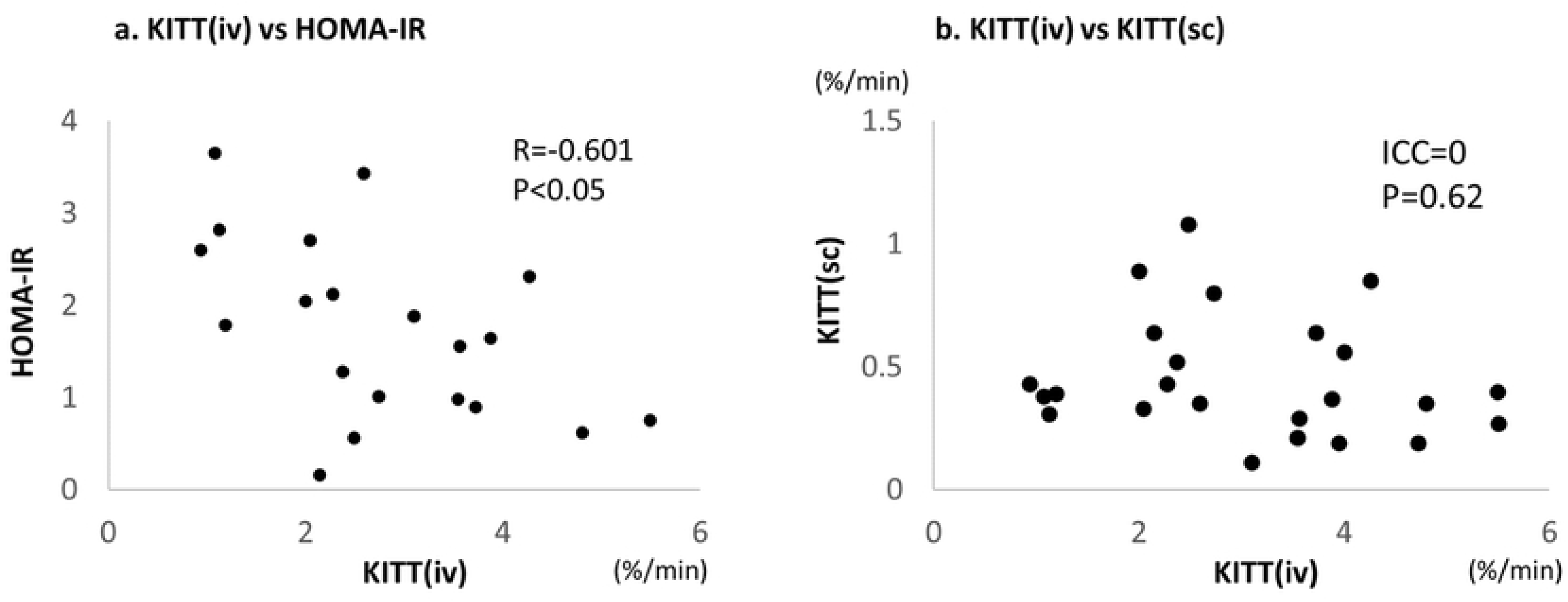
Relationship of K_ITT_(iv) with the HOMA-IR value and K_ITT_(sc). (a)K_ITT_(iv) was significantly correlated with the HOMA-IR value (r=−0.601, p<0.05). (b) K_ITT_(iv) was not correlated with K_ITT_(sc). KITT(iv), K index of the intravenous insulin tolerance test; HOMA-IR, homeostasis model assessment for insulin resistance; K_ITT_(sc), K index of the subcutaneous insulin tolerance test

**Fig 3.**
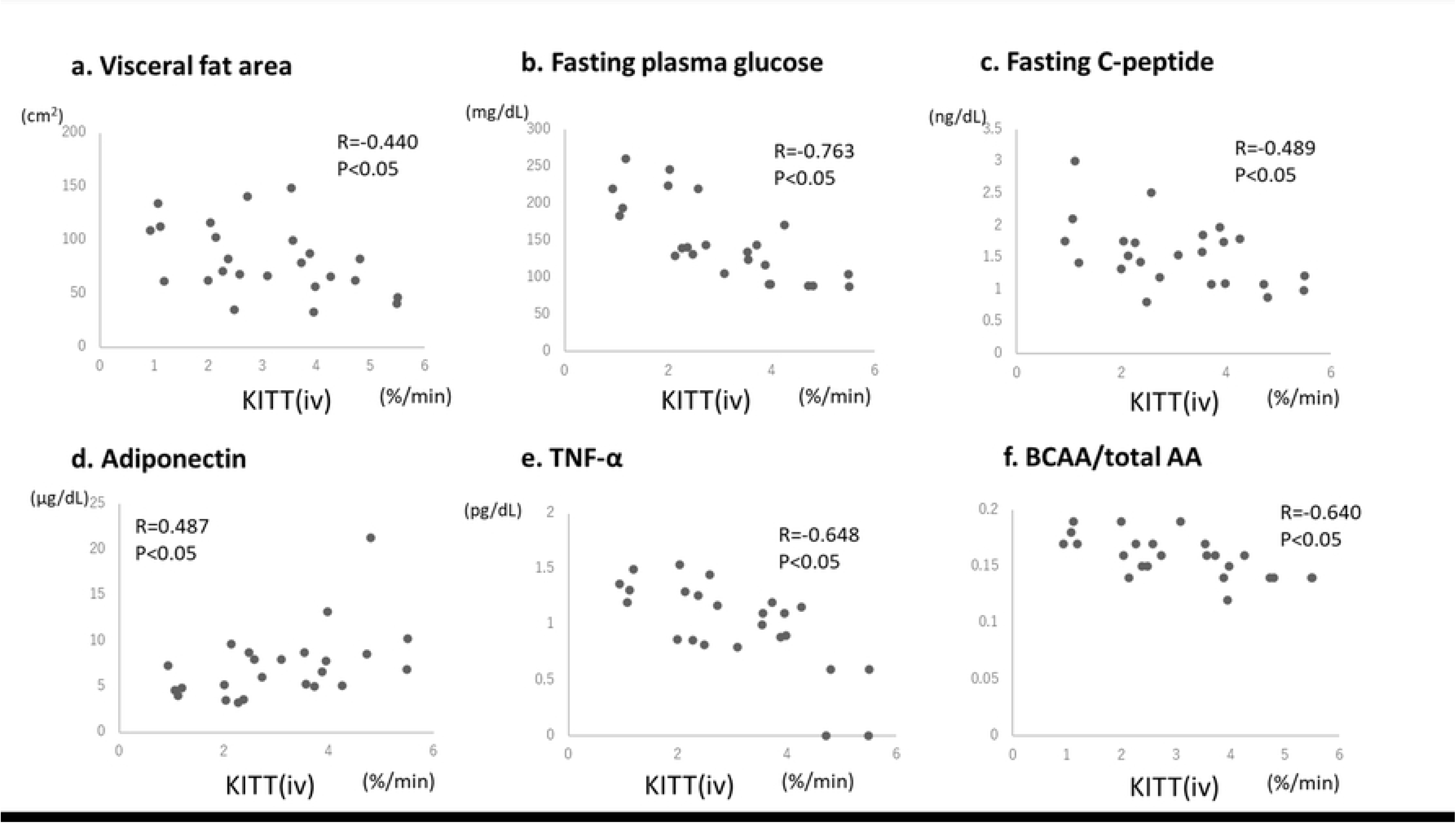
Relationship of K_ITT_(iv) with other parameters. (a)K_ITT_(iv) was negatively correlated with the visceral fat area (r=−0.444, p<0.05), (b)fasting plasma glucose (r=−0.673, p<0.05), (c)fasting serum C-peptide (r=−0.489, p<0.05), (e)TNF-α (r=−0.648, p<0.05), and (f)BCAA/total AA (r=−0.64, p<0.05). (d)It was positively correlated with the adiponectin level (r=0.487, p<0.05). K_ITT_(iv), K index of the intravenous insulin tolerance test; TNF-α, tumor necrosis factor-α; BCAA, branched-chain amino acid; AA, amino acid

In the 17 patients with T2DM, the mean total insulin dose used for glycemic control was 36.2±28.1 U/day, and the mean total insulin dose/body weight was 0.5±0.4 U/kg. The mean total insulin dose/body weight was negatively correlated with K_ITT_(iv) (r=−0.550, p<0.05), and it was calculated using the following regression equation (Fig 4a): total insulin dose/kg = 0.78–0.10× K_ITT_(iv). The mean total insulin dose/body weight was positively correlated with the HOMA-IR value (r=0.806, p<0.05), and it was calculated using the following regression equation (Fig 4b): total insulin dose/kg = 0.23+0.15× HOMA-IR value.

**Fig 4.**
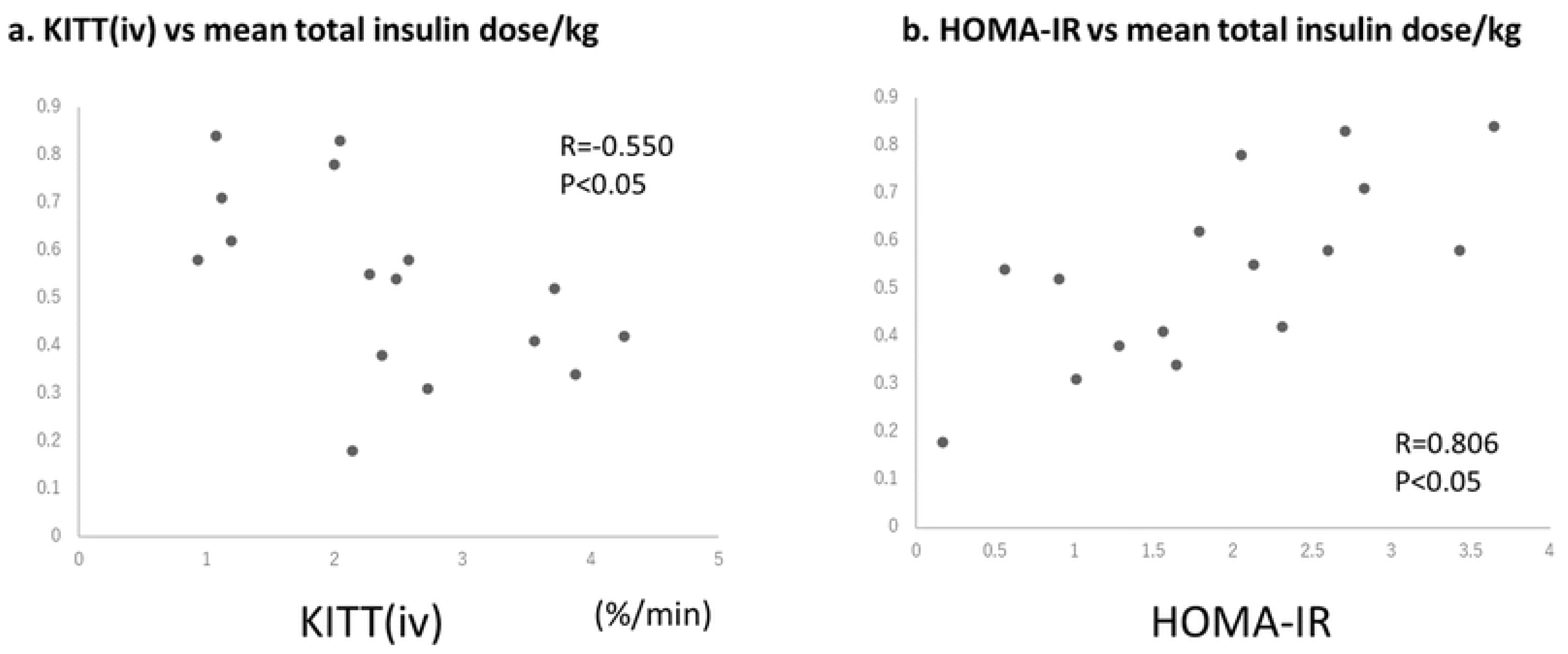
Relationship of K_ITT_(iv) and the HOMA-IR value with the mean total insulin dose/kg. (a)K_ITT_(iv) was significantly correlated with the mean total insulin dose per body weight (r=−0.550, p<0.05); this was estimated using with the following regression equation: total insulin dose/kg = 0.78–0.10 × K_ITT_(iv). (b)The HOMA-IR value was also significantly correlated with the mean total insulin dose per body weight (r=0.806, p<0.05); this was estimated using with the following regression equation: total insulin dose/kg = 0.23+0.15 × HOMA-IR. K_ITT_(iv), K index of the intravenous insulin tolerance test; HOMA-IR, homeostasis model assessment for insulin resistance

Seven and two adverse events occurred in participants undergoing the SITT and ITTsc, respectively. All adverse events were hypoglycemia. No significant difference was observed in the frequency of adverse events between participants undergoing the SITT and the ITTsc.

## Discussion

In the present study, we prospectively studied seven healthy controls and 14 patients with T2DM and evaluated their insulin resistance using K_ITT_(iv). The following four main results were obtained: 1) the K_ITT_(iv) of patients with T2DM was 2.5%±2.1%, which was significantly lower than that of the controls (4.5%±1.8%); 2) K_ITT_(iv) was significantly correlated with the HOMA-IR value but not with K_ITT_(sc); 3) K_ITT_(iv) was significantly correlated with VFA and the serum glucose, CPR, TNF-α, BCAA/total AA, and adiponectin concentrations; and 4) the total insulin dose required for glycemic control was correlated with K_ITT_(iv) and the HOMA-IR value, and it was estimated using an equation in patients with T2DM.

HOMA-IR values are widely used as indices of insulin resistance [13, 14]; however, their reliability decreases when the insulin secretion is reduced and hyperglycemia occurs [4]. In the present study, there was a significant correlation between the HOMA-IR value and K_ITT_(iv), suggesting its applicability as an index of insulin resistance even in a cohort including healthy volunteers and patients with poorly controlled T2DM. The significant correlation between the HOMA-IR value and K_ITT_(iv) may be attributed to the fact that the FPG level was not very high and the decrease in insulin secretion was only mild in our patients with T2DM. A previous study revealed that the HOMA-IR value is a useful index for determining insulin resistance at an FPG range of 80–170 mg/dL in obese Japanese patients with T2DM [4]. In fact, in this study, the mean FPG level of patients with T2DM was approximately 170 mg/L (Table 1), and the correlation observed between CPR and K_ITT_ supports the fact that the HOMA-IR value can be used as an effective index in this group (Fig 2b). In addition, the association between the two indices was observed even in the controls, which might have led to the significant correlation between the two indices in all study participants. Although the HOMA-IR data in this study were exclusive of patients on insulin therapies for diabetes, Okita et al. [10] reported that K_ITT_ in such patients can be a useful indicator of insulin resistance using the euglycemic clamp test.

We expected that K_ITT_(sc) would be correlated with K_ITT_(iv) because the pattern of glucose level changes, unlike the rate of action, in an individual may be similar between intravenous and subcutaneous routes of insulin administration. In addition, the accuracy of finger-prick tests for blood glucose measurement has substantially improved [15]. However, contrary to our expectations, there was no significant correlation between K_ITT_(iv) and K_ITT_(sc) (ICC=0). A possible explanation for this dissociation may be the instability of the insulin absorption rate due to differences in the skinfold thickness and subcutaneous blood flow [16, 17]. Exercise, smoking, and body position are also reported to affect insulin absorption [16, 17]. However, in this study, all patients were resting, had stopped smoking, and were sitting; thus, we considered these factors to have no effect. Previous studies [18-21] have compared glycemic control between intravenous and subcutaneous routes of insulin administration mainly in intensive care units; however, to our knowledge, there are no crossover studies comparing the two routes in the same individuals.

In this study, K_ITT_(iv) was correlated with various clinical parameters associated with obesity and insulin resistance, including VFA, adiponectin, tumor necrosis factor alpha (TNF-α), and branched chain amino acids (BCAA)/total AA. Obesity and visceral fat are closely associated with insulin resistance; thus, the significant correlation between KITT(iv) and the VFA was considered reasonable [10, 22]. Low adiponectin levels were also associated with an increased risk of insulin resistance [23], and TNF-α is a strong inhibitor of adiponectin promoter activity [24]. In non-obese, non-diabetic individuals, no significant correlation was observed between adipose tissue TNF-α mRNA content and K_ITT_(iv) [25]. However, when including patients with T2DM and a tendency for obesity, positive and negative correlations of K_ITT_(iv) with adiponectin and TNF-α, respectively, were observed in prior investigations similar to in our study[10]. In addition, in our study, K_ITT_(iv) was negatively associated with BCAAs/total AAs. This corroborates the findings of prior studies that revealed a significant association between insulin resistance and BCAAs [26, 27]. In fact, BCAAs were reported to accumulate under conditions of insulin resistance [28]; furthermore, a positive association of insulin resistance with valine, one of the three BCAAs, has been reported in Japanese healthy individuals [29].

To the best of our knowledge, this is the first report on a regression equation for calculating the insulin dose required for glycemic control based on K_ITT_(iv). It is useful for adjusting the insulin dose and avoiding hypoglycemic events. Notably, the correlation coefficient was better for the HOMA-IR value (r=0.806) than for K_ITT_(iv) (r=−0.550), suggesting that the HOMA-IR value is an easily applicable parameter that helps estimate the insulin dose required for glycemic control.

This study had several limitations. First, the sample size was small and the study was conducted at a single center. Second, there was significant heterogeneity among participants in terms of the age, sex, comorbidities, and treatment regimens (in patients with T2DM). The first and second limitations precluded robust statistical analyses. Finally, we did not verify the reproducibility and reliability of the equations for calculating the total insulin dose using the HOMA-IR value and K_ITT_(iv).

In conclusion, K_ITT_(iv) was associated with the HOMA-IR value and insulin resistance-related parameters (such as VFA, adiponectin, and BCAAs). K_ITT_(iv) is an index of insulin resistance that can be applied in clinical practice; along with HOMA-IR, it may be useful for efficiently adjusting insulin doses.

## Acknowledgements

None

## Data availability

The date presented in this manuscript are accessible and can be provided on reasonable request to the corresponding author.

## References

1. DeFronzo RA. Lilly lecture 1987. The triumvirate: beta-cell, muscle, liver. A collusion responsible for NIDDM. Diabetes. 1988;37: 667–687.

2. Godsland IF, Stevenson JC. Insulin resistance: syndrome or tendency? Lancet. 1995;346: 100–103.

3. Bonora E, Moghetti P, Zancanaro C, Cigolini M, Querena M, Cacciatori V, et al. Estimates of in vivo insulin action in man: comparison of insulin tolerance tests with euglycemic and hyperglycemic glucose clamp studies. J Clin Endocrinol Metab. 1989;68: 374–378.

4. Ono T, Shiga N, Taneda Y, Umemura S. The fasting-plasma glucose range in which insulin resistance measured by homeostasis model assessment correlates with euglycemic clamping. J Japan Diab Soc. 1999;42: 1005–1011.

5. Young RP, Critchley JA, Anderson PJ, Lau MS, Lee KK, Chan JC. The short insulin tolerance test: feasibility study using venous sampling. Diabet Med. 1996;13: 429–433.

6. Hirst S, Phillips DI, Vines SK, Clark PM, Hales CN. Reproducibility of the short insulin tolerance test. Diabet Med. 1993;10: 839–1842.

7. Akinmokun A, Selby PL, Ramaiya K, Alberti KG. The short insulin tolerance test for determination of insulin sensitivity: a comparison with the euglycaemic clamp. Diabet Med. 1992;9: 432–427.

8. Chen CC, Wang TY, Hsu SY, Chen RH, Chang CT, Chen SJ. Is the short insulin tolerance test safe and reproducible? Diabet Med. 1998;15: 924–927.

9. Graci S, Baratta R, Degano C, Luppa A, Vigneri R, Frittitta L, et al. The intravenous insulin tolerance test is an accurate method for screening a general population for insulin resistance and related abnormalities. J Endocrinol Invest. 1999;22: 472–475.

10. Okita K, Iwahashi H, Kozawa J, Okauchi Y, Funahashi T, Imagawa A, et al. Usefulness of the insulin tolerance test in patients with type 2 diabetes receiving insulin therapy. J Diabetes Investig. 2014;5: 305–312.

11. American Diabetes Association. Diagnosis and classification of diabetes mellitus. Diabetes Care. 2010;33: S62–S69.

12. Lundbaek K. Intravenous glucose tolerance as a tool in definition and diagnosis of diabetes mellitus. Br Med J. 1962;1: 1507–1513.

13. González-González JG, Violante-Cumpa JR, Zambrano-Lucio M, Burciaga-Jimenez E, Castillo-Morales PL, Garcia-Campa M, et al. HOMA-IR as a predictor of health outcomes in patients with metabolic risk factors: a systematic review and meta-analysis. High Blood Press Cardiovasc Prev. 2022;29: 547-564.

14. Tahapary DL, Pratisthita LB, Fitri NA, Marcella C, Wafa S, Kurniawan F, et al. Challenges in the diagnosis of insulin resistance: focusing on the role of HOMA-IR and Tryglyceride/glucose index. Diabetes Metab Syndr. 2022;16: 102581.

15. King F, Ahn D, Hsiao V, Porco T, Klonoff DC. A review of blood glucose monitor accuracy. Diabetes Technol Ther. 2018;20: 843–856.

16. Hildebrandt P. Subcutaneous absorption of insulin in insulin-dependent diabetic patients. Influence of species, physico-chemical properties of insulin and physiological factors. Dan Med Bull. 1991;38: 337–346.

17. Gradel AKJ, Porsgaard T, Lykkesfeldt J, Seested T, Gram-Nielsen S, Kristensen NR, et al. Factors affecting the absorption of subcutaneously administered insulin: effect on variability. J Diabetes Res. 2018;2018: 1205121.

18. Neff K, Donegan D, MacMahon J, O’Hanlon C, Keane N, Agha A, et al. Management of parenteral nutrition associated hyperglycaemia: a comparison of subcutaneous and intravenous insulin regimen. Ir Med J. 2014;107: 141–143.

19. Bodur HA, Saygili F, Saygili S, Doganay LH, Yesil S. Continuous infusion of subcutaneous compared to intravenous insulin for tight glycaemic control in medical intensive care unit patients. Anaesth Intensive Care. 2008;36: 520–527.

20. Furnary AP, Zerr KJ, Grunkemeier GL, Starr A. Continuous intravenous insulin infusion reduces the incidence of deep sternal wound infection in diabetic patients after cardiac surgical procedures. Ann Thorac Surg. 1999;67: 352–360.

21. Pezzarossa A, Taddei F, Cimicchi MC, Rossini E, Contini S, Bonora E, et al. Perioperative management of diabetic subjects. Subcutaneous versus intravenous insulin administration during glucose-potassium infusion. Diabetes Care. 1988;11: 52–58.

22. Kahn SE, Hull RL, Utzschneider KM. Mechanisms linking obesity to insulin resistance and type 2 diabetes. Nature. 2006;444: 840–846.

23. Moon HU, Ha KH, Han SJ, Kim HJ, Kim DJ. The association of adiponectin and visceral fat with insulin resistance and β-cell dysfunction. J Korean Med Sci. 2018;34: e7.

24. Maeda N, Takahashi M, Funahashi T, Kihara S, Nishizawa H, Kishida K, et al. PPARgamma ligands increase expression and plasma concentrations of adiponectin, an adipose-derived protein. Diabetes. 2001;50: 2094–2099.

25. Frittitta L, Youngren JF, Sbraccia P, D’Adamo M, Buongiorno A, Vigneri R, et al. Increased adipose tissue PC-1 protein content, but not tumour necrosis factor-alpha gene expression, is associated with a reduction of both whole body insulin sensitivity and insulin receptor tyrosine-kinase activity. Diabetologia. 1997;40: 282–289.

26. Allam-Ndoul B, Guénard F, Garneau V, Barbier O, Pérusse L, Vohl M. Associations between branched chain amino acid levels, obesity and cardiometabolic complications. Integr Obesity Diabetes. 2015;1: 157–162.

27. Elshorbagy A, Jernerén F, Basta M, Basta C, Turner C, Khaled M, et al. Amino acid changes during transition to a vegan diet supplemented with fish in healthy humans. Eur J Nutr. 2017;56: 1953–1962.

28. Lynch CJ, Adams SH. Branched-chain amino acids in metabolic signalling and insulin resistance. Nat Rev Endocrinol. 2014;10: 723–736.

29. Takashina C, Tsujino I, Watanabe T, Sakaue S, Ikeda D, Yamada A, et al. Associations among the plasma amino acid profile, obesity, and glucose metabolism in Japanese adults with normal glucose tolerance. Nutr Metab (Lond). 2016;13: 5

